# Characteristics of COVID-19 patients admitted to a tertiary care hospital in Pune, India, and cost-effective predictors of intensive care treatment requirement

**DOI:** 10.1101/2020.11.26.20239186

**Authors:** Urvi Bhooshan Shukla, Sharvari Rahul Shukla, Sachin Bhaskar Palve, Rajiv Chintaman Yeravdekar, Vijay Madhusoothan Natarajan, Pradeep Tiwari, Chittaranjan Sakerlal Yajnik

## Abstract

**Background:** Maharashtra is one of the worst affected states in this pandemic.^2^ As of 30th September, Maharashtra has in total 1.4 million cases with 38,000 deaths. Objective was to study associations of severity of disease and need for ICU treatment in COVID-19 patients.

**Methods:** A retrospective study of clinical course in 800 hospitalized COVID-19 patients, and a predictive model of need for ICU treatment. Eight hundred consecutive patients admitted with confirmed COVID-19 disease.

**Results:** Average age was 41 years, 16% were <20 years of age, 55% were male, 50% were asymptomatic and 16% had at least one comorbidity. Using MoHFW India severity guidelines, 73% patients had mild, 6% moderate and 20% severe disease. Severity was associated with higher age, symptomatic presentation, elevated neutrophil and reduced lymphocyte counts and elevated inflammatory markers. Seventy-seven patients needed ICU treatment: they were older (56 years), more symptomatic and had lower SpO2 and abnormal chest X-ray and deranged hematology and biochemistry at admission. A model trained on the first 500 patients, using above variables predicted need for ICU treatment with sensitivity 80%, specificity 88% in subsequent 300 patients; exclusion of expensive laboratory tests did not affect accuracy.

**Conclusion:** In the early phase of COVID- 19 epidemic, a significant proportion of hospitalized patients were young and asymptomatic. Need for ICU treatment was predicted by simple measures including higher age, symptomatic onset, low SpO2 and abnormal chest X-ray. We propose a cost-effective model for referring patients for treatment at specialized COVID-19 hospitals.

**Key Messages:**

- Of 800 patients, 73% had mild, 6% moderate and 20% had severe disease.
- Seventy-seven patients (9.6%) required ICU treatment, 25 (3%) died.
- ICU treatment was predicted by higher age, more symptomatic presentation, lower SpO2 and pneumonia on chest X-ray at admission.
- A machine learning model features in first 500 patients accurately predicted ICU treatment in subsequent 300 patients.
- A good clinical protocol, SpO2 and chest X-ray are adequate to predict and triage COVID-19 patients for hospital admissions in resource poor environments.

## Introduction

India reported its first case of COVID-19 on 30th January, 2020 in Kerala.^1^ As of 30^th^ September 2020, according to the Ministry of Health and Family Welfare (MoHFW) more than 6 million COVID-19 cases have been reported in India, with 95,00 deaths. India is now 2nd only to United States in the number of cases.^2^

Maharashtra is one of the worst affected states in this pandemic.^2^ As of 30th September, Maharashtra has in total 1.4 million cases with 38,000 deaths. COVID-19 cases in Pune have exceeded those in Mumbai. There is considerable load on the health care system to accommodate COVID-19 patients which is met by opening specialized centers for mild patients. Guidelines for triaging patients for hospitalization who may need subsequent ICU treatment will be useful.

Symbiosis University Hospital and Research Center is a state of the art hospital. The management offered 500 isolation and 30 ICU beds for COVID-19 patients to the local authorities on 24th March 2020.

We narrate our experience of managing the initial 800 consecutive patients admitted with confirmed COVID-19 positive status. We described the demographic, clinical and biochemical characteristics, and developed a model of predictors for ICU admission.

## Methods

COVID-19 patient admissions began on 8^th^ April 2020, patients were referred by the Pune Municipal Corporation with a positive RT-PCR^3^ test for SARS-CoV2 done at the National Institute of Virology, Pune. The test was advised for symptoms suggestive of COVID-19 or because of close contact with COVID-19 patients.

All patients were screened at admission for severity of the disease by the MoHFW Clinical Guidelines for the management of COVID-19, dated 30^th^ March 2020 (Table-S1).^4^ Screening included: demographic details, history of symptoms, comorbidities and their medication, clinical examination including respiratory rate, blood pressure, pulse oximetry (SpO2) and chest X-ray. Clinically stable patients with SpO2 >94% on room air and without severe radiographic abnormality were classified as mild. Patients with lung infiltrates on chest X-ray were classified as having moderate disease if SpO2 was 90-94%, and severe if SpO2 was <90%.^4^

A venous blood sample was collected for following laboratory measurements: Complete blood count (CBC) with absolute neutrophil count (ANC), absolute lymphocyte count (ALC), liver and kidney tests, serum C-reactive protein (CRP), serum ferritin (as an acute phase reactant). Patients with comorbidities had appropriate additional tests (glucose, HbA1c etc) (Table-S2).

A baseline ECG was recorded for any abnormalities and specifically for QTc, in anticipation of hydroxychloroquine (HCQ, 400mg BD on day 1, and 200mg BD x 4 days) treatment which was not given for those with prolonged QTc, G6PD deficiency and those younger than 16 years of age. HCQ treatment was not prescribed for those patients admitted from June 2020 based on updated evidence on its efficacy in the treatment of COVID-19. Pre-admission treatment for co-morbidities was continued or adjusted as necessary.

Laboratory tests were repeated as necessary and usually on day 5 for stable patients (Table-S2). Those who showed deterioration in clinical state, fall in SpO2 or worsening chest X-ray were transferred to the Intensive Care Unit (ICU) for necessary treatment. Patients with a stable course were discharged between days 15-18 after two consecutive negative tests for SARS CoV-2 Ag. In these initial 800 patients there were no height and weight measurements during admission. We were able to obtain this data during a telephonic follow up 6 weeks after discharge (n=219).

All patients signed a written informed consent at the time of admission which permitted use of anonymized data for research. The Institutional Research Committee (IRC) of Symbiosis Medical College for Women gave necessary approvals.

### Statistical methods

We have presented data of clinical and laboratory characteristics of the initial 800 patients by sex, severity of disease and for those admitted to ICU. We have also shown data for those who were asymptomatic at admission. For statistical analysis, variables with skewed distributions were log-transformed to satisfy assumptions of normality. Differences in clinical and biochemical characteristics between groups of patients were tested by ANOVA adjusting for age and sex or by Chi-square test. Significant associations generated in this analysis were used as predictors to build a multivariate logistic regression model to test independent associations with the outcome of need for ICU treatment. These are presented as ROC curves. Analyses were carried out using IBM SPSS Statistics for Windows, version 23.0 (IBM Corp., Armonk, N.Y., USA).

A Random Forest model ^5^ was also built to predict ICU admission requirement using significant clinical and laboratory features from the above analysis. The model was trained on the first 500 patients and tested on subsequent 300 patients for its accuracy which is reported as sensitivity, specificity, positive predictive value (PPV) and negative predictive value (NPV). The data on subsequent 300 patients was shared for validation only after the model was available on the first 500 patients. We also generated models for a limited number of predictors which are likely to be available in a ‘primary care ‘set-up by excluding the more expensive and specialized measurements. The model was built with 10,000 trees using randomForest package^6^ in R statistical programming language.^7^

## Results

Of the initial 800 consecutive COVID-19 patients admitted to Symbiosis University Hospital between 8^th^ April to 15th June, 402 patients were tested for SARS-CoV2 because they had suggestive symptoms and others because of close contact with a case (Fig 1). There were 440 males and 360 females. Our cohort of COVID-19 patients is relatively young: 49% of patients were below 40 years of age (16% less than 20 years), only a fifth (n=155) were beyond 60 years of age. Majority of patients belonged to lower and lower-middle socio-economic classes (Table-S3).

**Figure 1:**
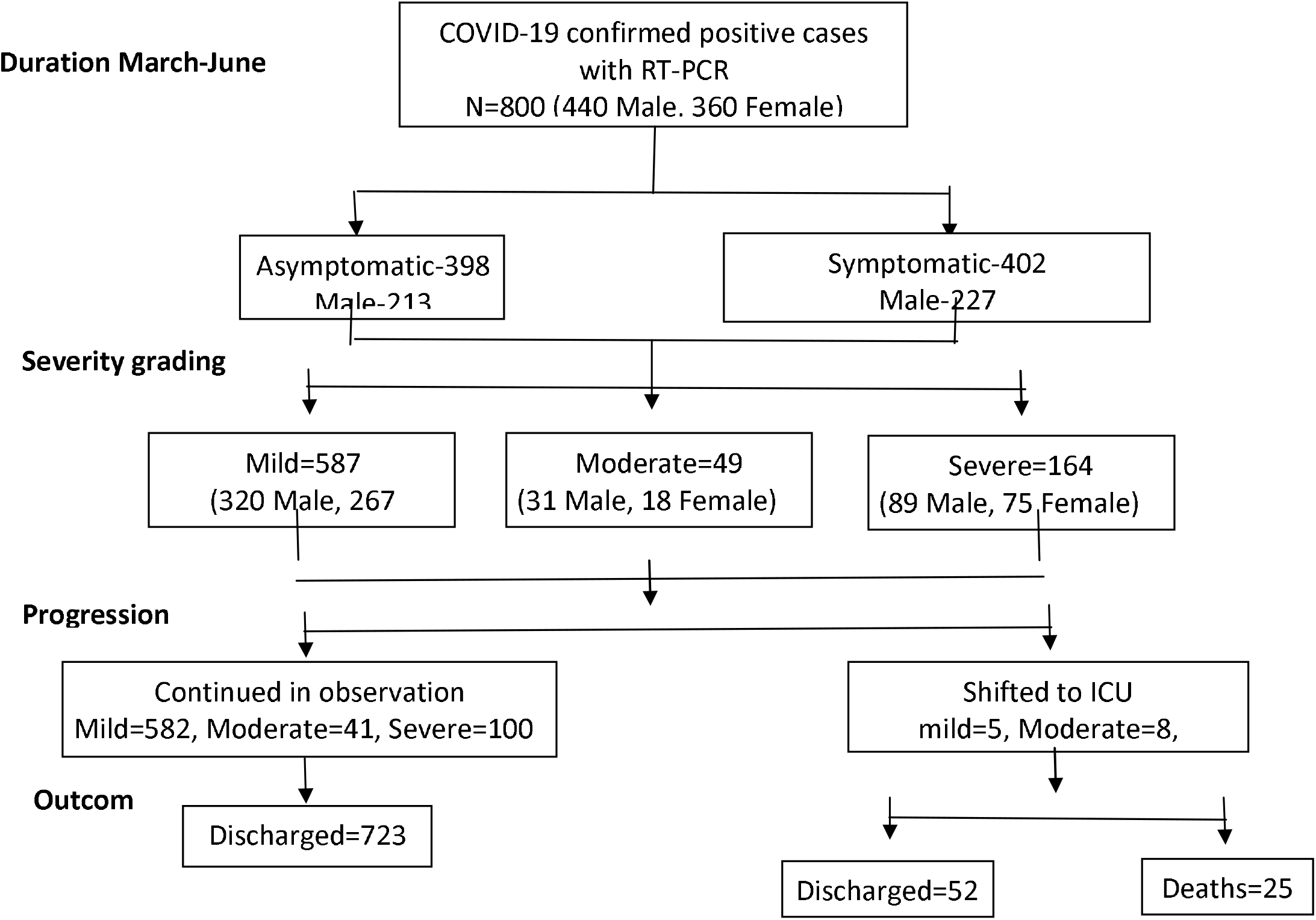
Flowchart of study participants

At admission (Table-S3) 50% of patients had no symptoms, 31% had one symptom, 14% had two symptoms and only 6% had more than two symptoms. The commonest symptoms were fever (26%) and cough (26%); other symptoms included sore throat (13%) and breathlessness (11%). The average duration of symptoms before hospitalization was 3.4 days. Diabetes (17%) and hypertension (15%) were the commonest comorbidities which were more common in females. None of the patients had a previous diagnosis of tuberculosis, HIV or other immunocompromised state. We were able to calculate BMI in 219 patients from self-reported height and weight data to be 25.0 ± 6.0 kg/m^2^ (age 45.4 ± 16.8 years, 52% males).

Asymptomatic patients were younger (38.9 vs 44.7 years), less likely to have comorbidities (23 vs 28%), and had lower prevalence of moderate and severe disease (15% vs 39% respectively) compared to those symptomatic at the time of hospital admission. Only 3% of asymptomatic patients needed ICU treatment and 4 died compared to 16% ICU admissions and 21 deaths in symptomatic group. When comparing laboratory tests, lymphopenia was more common in symptomatic and severely ill patients compared to asymptomatic patients, and inflammatory markers were also higher. As a group, mild and asymptomatic patients had near normal biochemical tests, normal chest X-ray (95%) that remained normal over the course of illness. Details are provided in Table-S5.

Table-1 describes characteristics of patients by the clinical severity at admission. Out of 800 patients, 587 (73%) had mild disease, 49 (6%) moderate and 164 (21%) severe disease. Twenty-four severely ill patients needed immediate ICU admission (for hypoxemic respiratory failure) (Table-S4), others were admitted in the isolation wards. Moderate and severely ill patients were older than those with mild disease, had larger number of symptoms at presentation and for longer duration. Twenty-four percent of patients with severe disease had more than one comorbidity (35% had diabetes and 31% had hypertension). Expectedly, SpO2 was lower in moderate and severe cases. Chest X-Ray was normal in 72%, and showed unilateral or bilateral infiltrates in 28% patients which were mostly seen in moderate and severe groups. Pulse rate and blood pressure were similar across groups.

**Table 1:** Demographic, clinical characteristics and comorbidity of patients by severity grading

| **Characteristic** | **Mild**<br>**n (%)** | **Moderate**<br>**n (%)** | **Severe**<br>**n (%)** | **p** | **p1** |
| --- | --- | --- | --- | --- | --- |
| N | 587 (73.4) | 49 (6.1) | 164 (20.5) | -- | -- |
| Age (mean, SD) | 37.3 (19.1) | 52.7 (19.1) | 54.9 (15.5) | <.001 | -- |
| **Age categories (y)\*** |  |  |  |  |  |
| < 10 | 50 (8.5) | 2 (4.1) | 1 (0.6) | <.001 | -- |
| 10-20 | 71 (12.1) | 1 (2.0) | 1 (0.6) |  |  |
| 20-40 | 231 (39.4) | 10 (20.4) | 28 (17.1) |  |  |
| 40-60 | 156 (26.6) | 18 (36.7) | 76 (46.3) |  |  |
| >60 | 79 (13.5) | 18 (36.7) | 58 (35.4) |  |  |
| **Sex\*** |  |  |  |  |  |
| Male | 320 (54.5) | 31 (63.3) | 89 (54.3) | .49 | -- |
| Female | 267 (45.5) | 18 (36.7) | 75 (45.7) |  |  |
| **Number of**<br>**symptoms at**<br>**admission\*** |  |  |  |  |  |
| Nil | 340 (57.9) | 17 (34.7) | 41 (25.0) | <.001 | -- |
| 1 | 177 (30.2) | 18 (36.7) | 52 (31.7) |  |  |
| 2 | 58 (9.9) | 10 (20.4) | 41 (25.0) |  |  |
| >2 | 12 (2.0) | 4 (8.2) | 30 (18.3) |  |  |
| **Symptoms\*** |  |  |  |  |  |
| Fever | 130 (22.1) | 14 (28.6) | 61 (37.2) | <.001 | -- |
| Cold-Cough | 118 (20.1) | 21(42.9) | 71 (43.3) | <.001 | -- |
| Sore Throat | 66 (11.2) | 7 (14.3) | 32 (19.5) | .02 | -- |
| Breathless | 16 (2.7) | 9 (18.4) | 65 (39.6) | <.001 | -- |
| Duration of symptoms (days) | 2.6 (1.4) | 3.2 (1.9) | 6.3 (11.4) | <.01 | -- |
| **Comorbidity\*** |  |  |  |  |  |
| T2DM | 67 (11.4) | 11 (22.4) | 58 (35.4) | <.001 | -- |
| Hypertension | 63 (10.7) | 6 (12.2) | 50 (30.5) | <.001 | -- |
| IHD/CABG | 10 (1.7) | 2 (4.1) | 11 (6.7) | <.01 | -- |
| Asthma/COPD | 3 (0.5) | -- | 10 (6.1) | -- | -- |
| **Number of comorbidities\*** |  |  |  |  |  |
| Nil | 473 (80.6) | 35 (71.4) | 87 (53.0) | <.001 | -- |
| One | 82 (14.0) | 10 (20.4) | 38 (23.2) |  |  |
| More than one | 32 (5.5) | 4 (8.2) | 39 (23.8) |  |  |
| Blood pressure<br>(mmHg) |  |  |  |  |  |
| Systolic BP | 120 (110-130) | 122 (116-130) | 127 (112-137) | <.001 | .69 |
| Diastolic BP | 70 (70-80) | 80 (71-90) | 75 (70-80) | .12 | .03 |
| Heart rate | 78 (72-84) | 76 (74-82) | 78 (71-86) | .94 | .22 |
| **Biochemical**<br>**characteristics** |  |  |  |  |  |
| **Day 1 (n=778)** |  |  |  |  |  |
| Hemoglobin (g/L) | 127 (114-141) | 126 (106-139) | 126 (115-139) | .99 | .32 |
| TLC ($\times 10^3$/cmm) | 6.1 (4.7-7.6) | 5.8 (4.8-7.5) | 6.5 (4.9-8.7) | <.01 | .02 |
| Platelet count<br>($\times 10^5$/cmm) | 242 (197-299) | 221 (155-320) | 232 (184-298) | .92 | .17 |
| ANC ($\times 10^3$/cmm) | 3.2 (2.3-4.4) | 3.6 (3.0-4.9) | 4.5 (3.0-6.8) | <.001 | <.001 |
| ALC ($\times 10^3$/cmm) | 1.8 (1.3-2.4) | 1.4 (0.95-2.0) | 1.1 (0.72-1.6) | <.001 | <.001 |
| ANC/ALC | 1.78 (1.23-2.67) | 2.46 (1.76-3.94) | 3.73 (2.10-7.80) | <.001 | <.001 |
| CRP (nmol/l) | 14.19 (5.61-47.02) | 180.00(36.38-686.77) | 413.72 (70.19-928.87) | <.001 | <.001 |
| S. Ferritin (pmol/l) | 126.73 (38.42-278.62) | 473.21 (162.23-1165.968) | 627.81 (239.97-1456.05) | <.001 | <.001 |
| PT (sec) | 11.2 (10.8-11.9) | 11.1 (10.6-11.9) | 11.2 (10.6-12.6) | .05 | .09 |
| INR | 1.01 (0.97-1.08) | 1.01 (0.96-1.08) | 1.02 (0.96-1.14) | .05 | .12 |
| **ECG (n=356)\*** |  |  |  |  |  |
| Normal | 237 (86.5) | 20 (95.2) | 43 (70.5) | <.01 | -- |
| Abnormal | 37 (13.5) | 1 (4.8) | 18 (29.5) |  |  |
| **Chest X-ray \*** |  |  |  |  |  |
| Normal | 558 (95.1) | 14 (28.6) | 5 (3.0) | -- | -- |
| Unilateral infiltrates | 29 (4.9) | 35 (71.4) | 3 (1.8) |  |  |
| Bilateral infiltrates | -- | -- | 156 (95.1) |  |  |
| **Day 5 (n=581)** |  |  |  |  |  |
| Hemoglobin (g/L) | 127 (112-141) | 124 (108-136) | 12.4 (111-137) | .21 | .17 |
| TLC (x10<sup>3</sup>/cmm) | 6.4 (5.2-7.8) | 6.8 (5.4-7.5) | 7.5 (5.5-11.0) | .77 | .02 |
| Platelet count (x<br>10<sup>5</sup>/cmm) | 259 (207-313) | 291 (191.7-397.2) | 296.5 (220.7-369.2) | <.01 | <.01 |
| ANC (x10<sup>3</sup>/cmm) | 3.1 (2.3-4.1) | 4.1 (3.2-4.9) | 5.1 (2.8-8.7) | <.001 | <.001 |
| ALC (x10<sup>3</sup>/cmm) | 2.2 (1.7-2.8) | 1.3 (0.97-2.05) | 1.1 (0.70-1.72) | <.001 | <.001 |
| ANC/ALC | 1.38 (1.0-2.02) | 2.73 (1.70-6.19) | 4.18 (1.89-11.31) | <.001 | <.001 |
| CRP (nmol/L) | 15.04 (6.09-50.28) | 145.71 (27.71-490.39) | 177.52(36.38-696.20) | <.001 | <.001 |
| S. Ferritin (pmol/L) | 140.66 (54.37-309.41) | 532.31 (188.07-1014.97) | 587.14 (315.25-1517.84) | <.001 | <.001 |
| Creatinine (μmol/L) | 53.37 (45.75-68.62) | 61.00 (53.37-76.25) | 68.62 (53.37-83.87) | .85 | .28 |
| **Outcome\***<br>**ICU Transfer** | 5 (0.9) | 8 (16.3) | 64 (39.0) | <.001 | -- |
| Death | 2 (0.3) | 1 (2.0) | 22 (13.4) | -- | -- |

Note: values are Median (IQR) and p by ANOVA, p1 adjusted for age and sex. \* number (%) and p by $\chi^2$ -test.
T2DM- Type-2 Diabetes Mellitus, IHD-Ischemic heart disease, CABG-Coronary artery bypass graft , COPD- Chronic obstructive pulmonary disease, TLC-Total Leucocyte count, ANC- Absolute Neutrophil Count, ALC- Absolute lymphocyte count, CRP- C-Reactive protein, PT- Prothrombin time, INR- International normalized ratio.

Hemoglobin, total leucocyte count and platelet count were similar across three severity groups. There was increasing neutrophilia and lymphopenia with increasing severity which reflected in increasing ANC/ALC ratio. Markers of inflammation i.e., serum C-reactive protein (CRP) and serum ferritin progressively increased with severity of the disease. The haematological and inflammatory markers remained stable and near normal in mild cases (on second measurement 3-5 days after admission) but continued to be elevated or deteriorated in severe patients.

Table-S3 shows demographic and clinical characteristics by sex. Females had similar age, symptoms and severity grading but had higher rate of co-morbidities compared to males. Mortality was higher in males.

HCQ was prescribed to 395 patients. It was stopped in 12 patients who developed QTc prolongation on ECG. Seventy-seven patients also received Azithromycin and 86 Oseltamivir, based on treating physician’s preference.

A total of 77 patients needed ICU treatment (Table-2) (24 directly admitted, 53 shifted from isolation ward due to clinical deterioration). These patients had a mean age of 57 years (min 25 and max 90 years) and 44 were males. Majority of these patients were symptomatic at admission, had longer duration of symptoms and about half of these patients had at least one comorbidity (29 diabetes and 26 hypertension). They had higher ANC (median 5.3 vs 3.2), lower ALC (median 1.0 vs 1.8), and higher ANC/ALC ratio (median 5.3 vs 1.9) compared to those who did not require ICU treatment. Similarly, CRP and serum ferritin concentrations were also higher Ten of these patients had cardiac arrhythmias, 37 required invasive mechanical ventilation, 12 had acute kidney injury (KDIGO^8^ stage 1 and above) and 25 developed circulatory failure (Table-S4). Twenty-five patients died in the ICU: four because of refractory hypoxemia and 19 due to multi-organ failure, 2 due to sudden cardiac death. Patients who died were older (mean age 64.7, min 36-max 90 years) compared to those who survived, and predominantly male 18/25. Fifty-two patients were discharged from ICU; the average duration of ICU stay was 12 days.

**Table 2:** Demographic, clinical characteristics and comorbidity of patients by ICU admission

| **Characteristic** | **ICU admission** |  | **p** | **p1** |
| --- | --- | --- | --- | --- |
|  | **No**<br>**n (%)** | **Yes**<br>**n (%)** |  |  |
| N | 723 (90.4) | 77 (9.6) | -- | -- |
| Age (mean, SD)* | 40.2 (19.5) | 57.4 (15.9) | <.001 | -- |
| **Age categories (y)\*** |  |  |  |  |
| < 10 | 53 (7.3) | -- | <.001 | -- |
| 10-20 | 72 (10.0) | 1 (1.3) |  |  |
| 20-40 | 257 (35.5) | 12 (15.6) |  |  |
| 40-60 | 217 (30.0) | 33 (42.9) |  |  |
| >60 | 124 (17.2) | 31 (40.3) |  |  |
| **Sex\*** |  |  |  |  |
| Male | 396 (54.8) | 44 (57.1) | .69 | -- |
| Female | 327 (45.2) | 33 (42.9) |  |  |
| **Number of symptoms at admission\*** |  |  |  |  |
| Nil | 387 (53.5) | 11 (14.3) | <.001 | -- |
| 1 | 222 (30.7) | 25 (32.5) |  |  |
| 2 | 84 (11.6) | 25 (32.5) |  |  |
| >2 | 30 (4.1) | 16 (20.8) |  |  |
| **Symptoms\*** |  |  |  |  |
| Fever | 171 (23.7) | 34 (44.2) | <.001 | -- |
| Cold-Cough | 171 (23.7) | 39 (50.6) | <.001 | -- |
| Sore Throat | 94 (13.0) | 11 (14.3) | .89 | -- |
| Breathless | 49 (6.8) | 41 (53.2) | <.001 | -- |
| Duration of symptoms (days) | 2.6 (1.5) | 7 (12.6) | <.01 | -- |
| **Comorbidity\*** |  |  |  |  |
| T2DM | 107 (14.8) | 29 (37.7) | <.001 | -- |
| Hypertension | 93 (12.9) | 26 (33.8) | <.001 | -- |
| IHD/CABG | 15 (2.1) | 8 (10.4) | <.001 | -- |
| Asthma/COPD | 5 (0.7) | 8 (10.4) | <.001 | -- |
| **Number of comorbidity\*** |  |  |  |  |
| Nil | 558 (77.2) | 37 (48.1) | <.001 | -- |
| One | 109 (15.1) | 21 (27.3) |  |  |
| More than one | 56 (7.7) | 19 (24.7) |  |  |
| **Severity grading at admission\*** |  |  |  |  |
| Mild | 582 (80.5) | 5 (6.5) | <.001 | -- |
| Moderate | 41 (5.7) | 8 (10.4) |  |  |
| Severe | 100 (13.8) | 64 (83.1) |  |  |
| Blood pressure (mmHg) |  |  |  |  |
| Systolic BP | 120 (110-130) | 130 (110-150) | <.001 | .52 |
| Diastolic BP | 75 (70-80) | 74 (70-81) | .59 | .05 |
| Heart rate | 78 (72-83) | 80 (73-90) | .08 | <.01 |
| **Biochemical characteristics** |  |  |  |  |
| **Day 1 (778)** |  |  |  |  |
| Hemoglobin (g/L) | 127 (114-141) | 121 (109-137) | .07 | <.01 |
| TLC ($\times 10^3/\text{cmm}$) | 6.1 (4.8-7.7) | 6.8 (4.9-9.3) | .10 | .03 |
| Platelet count ($\times 10^5/\text{cmm}$) | 241 (194-300) | 222 (165-297) | .46 | .99 |
| ANC ($\times 10^3/\text{cmm}$) | 3.3 (2.4-4.6) | 5.3 (3.1-8.3) | <.001 | <.001 |
| ALC ($\times 10^3/\text{cmm}$) | 1.80 (1.20-2.30) | 1.0 (0.60-1.40) | <.001 | <.001 |
| ANC/ALC | 1.89 (1.29-2.87) | 5.26 (2.53-10.07) | <.001 | <.001 |
| CRP (nmol/L) | 19.04 (6.57-99.24) | 854.30 (314.29-1189.548) | <.001 | <.001 |
| S. Ferritin (pmol/L) | 156.61 (53.47-377.04) | 915.2(443.1-2054.65) | <.001 | <.001 |
| PT (sec) | 11.2 (10.7-11.9) | 11.2 (10.7-12.8) | <.01 | .008 |
| INR | 1.01 (0.97-1.08) | 1.02 (0.96-1.14) | <.01 | <.01 |
| **ECG (n=356)\*** |  |  |  |  |
| Normal | 275 (86.5) | 25 (65.8) | <.01 | -- |
| Abnormal | 43 (13.5) | 13 (34.2) |  |  |
| **Chest X-ray** |  |  |  |  |
| Normal | 275 (63.0) | 3 (7.5) | <.001 | -- |
| Abnormal | 161 (37.0) | 37 (92.5) |  |  |
| **Day 5 (n=581)** |  |  |  |  |
| Hemoglobin (g/L) | 126 (112-140) | 123 (107-138) | .06 | .04 |
| TLC (x10<sup>3</sup>/cmm) | 6.4 (5.2-7.8) | 6.7 (6.4-13.6) | .05 | <.001 |
| Platelet count (x 10<sup>5</sup>/cmm) | 264 (208-324) | 275 (212-369) | .29 | .89 |
| ANC (x10<sup>3</sup>/cmm) | 3.2 (2.3-4.3) | 6.4 (4.3-10.9) | <.001 | <.001 |
| ALC (x10<sup>3</sup>/cmm) | 2.10 (1.50-2.70) | 0.80 (.40-1.35) | <.001 | <.001 |
| ANC/ALC | 1.5 (1.1-2.3) | 8.2 (3.1-25.1) | <.001 | <.001 |
| CRP (nmol/L) | 18.09 (6.76-71.14) | 465.72 (117.14-913.35) | <.001 | <.001 |
| S. Ferritin (pmol/L) | 175.26 (65.16-424.68) | 984.18 (377.49-1804.34) | <.001 | <.001 |
| Creatinine (μmol/L) | 53.37 (45.75-68.62) | 68.62 (57.18-115.13) | .72 | <.01 |
| **Outcome\*** |  |  |  |  |
| Death | 1 (0.1) | 24 (31.2) | -- | -- |

Note: values are Median (IQR) and p by ANOVA, p1 adjusted for age and sex. \* number (%) and p by $\chi^2$ -test.

We found that the following factors were significantly related to ICU admission on univariate analysis: higher age, larger number of symptoms at diagnosis, more than 1 co-morbidity, abnormal chest X-ray, low SpO2 (< 94%), higher ANC/ALC ratio, CRP and Serum ferritin concentrations.

Table-3 describes the multivariate logistic regression to predict ICU treatment. Significant predictors were: higher age (OR 1.05, 95% CI 1.04 to 1.06), symptomatic presentation (OR 6.13, 95% CI 3.14 to 11.96), abnormal chest X-ray (OR 20.14, 95% CI 8.79 to 46.16) and low SpO2 < 93% (OR 12.67, 95% CI 6.36 to 25.23). Two indicators of lung involvement (low SpO2 and abnormal chest X-ray) had an overriding prediction. The laboratory tests (ANC/ALC ratio, CRP, S. Ferritin) were significantly related but did not make an additional contribution to the prediction (Fig 2). The overall prediction was high (AUC 0.951), and the model suggests that simple clinical, radiological and SpO2 predict requirement for ICU treatment quite reliably. Of the laboratory tests, the simple hematological test of CBC made an equal contribution compared to the costlier markers of inflammation. HCQ treatment did not reduce the need for ICU treatment.^9^

**Table 3:** Multivariate model for predictors of ICU admission.

|  |  | **Model for ICU admission** |  |  |  |  |
| --- | --- | --- | --- | --- | --- | --- |
|  | Variables | Std beta | Lower CI | Upper CI | P | AUC |
| Model 1 | **Age (y)** | 1.051 | 1.036 | 1.067 | <.001 |  |
|  | **Sex** |  |  |  |  |  |
|  | Female | 1 |  |  |  |  |
|  | Male | 1.202 | 0.733 | 1.972 | .47 | .75 |
| Model 2 | **+ Symptoms** |  |  |  |  |  |
|  | No | 1 |  |  |  |  |
|  | Yes | 6.132 | 3.142 | 11.968 | <.001 | .81 |
| Model 3 | **+ Co-morbidity** |  |  |  |  |  |
|  | No | 1 |  |  |  |  |
|  | Yes | 1.862 | 1.071 | 3.238 | .03 | .82 |
| Model 4 | **+ Chest X-ray** |  |  |  |  |  |
|  | Normal | 1 |  |  |  |  |
|  | Abnormal | 20.141 | 8.787 | 46.163 | <.001 | .91 |
| Model 5 | **+ SpO2** |  |  |  |  |  |
|  | >93% | 1 |  |  |  |  |
|  | <93% | 12.671 | 6.362 | 25.235 | <.001 | .95 |
| Model 6-1 | + ANC/ALC ratio | 1.449 | 0.945 | 2.221 | <.001 | .95 |
| Model 6-2 | + C-reactive protein | 1.503 | 1.183 | 1.910 | .001 | .95 |
| Model 6-3 | + S. Ferritin | 1.459 | 1.088 | 1.955 | .012 | .95 |

Note: All models adjusted for age and sex. Individuals independent variables are added in model stepwise. ALC/ALC ratio, CRP and S Ferritin added separately after Model 5.

**Fig 2:**
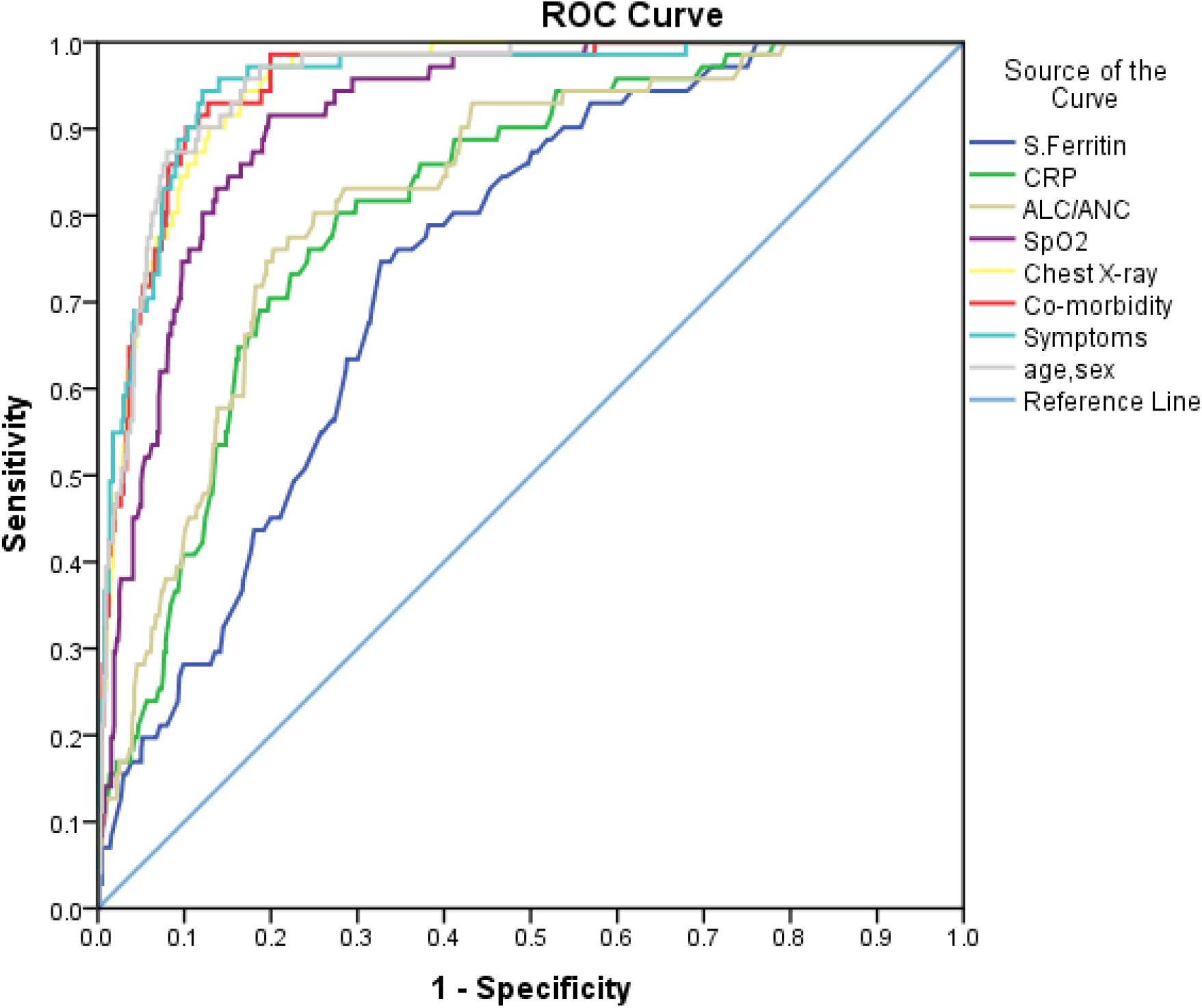
The ROC curve for predictors of ICU admission.

A Random forest model was constructed on initial 500 patients using sex and eight features found to be significant in univariate logistic regression. The model was able to predict ICU admission with high accuracy on subsequent 300 patients (sensitivity 80%, specificity 88%, PPV 47%, NPV 97%). CRP, SpO2 and chest X-ray were the top three important predictors. A model without CRP and ferritin was comparable (sensitivity 77%, specificity 86%, PPV 43% and NPV 96%), further excluding younger patients (<20 years of age) only marginally improved the accuracy. (sensitivity 87%, specificity 86%, PPV 46% and NPV 98%) (Table-4)

**Table 4:**
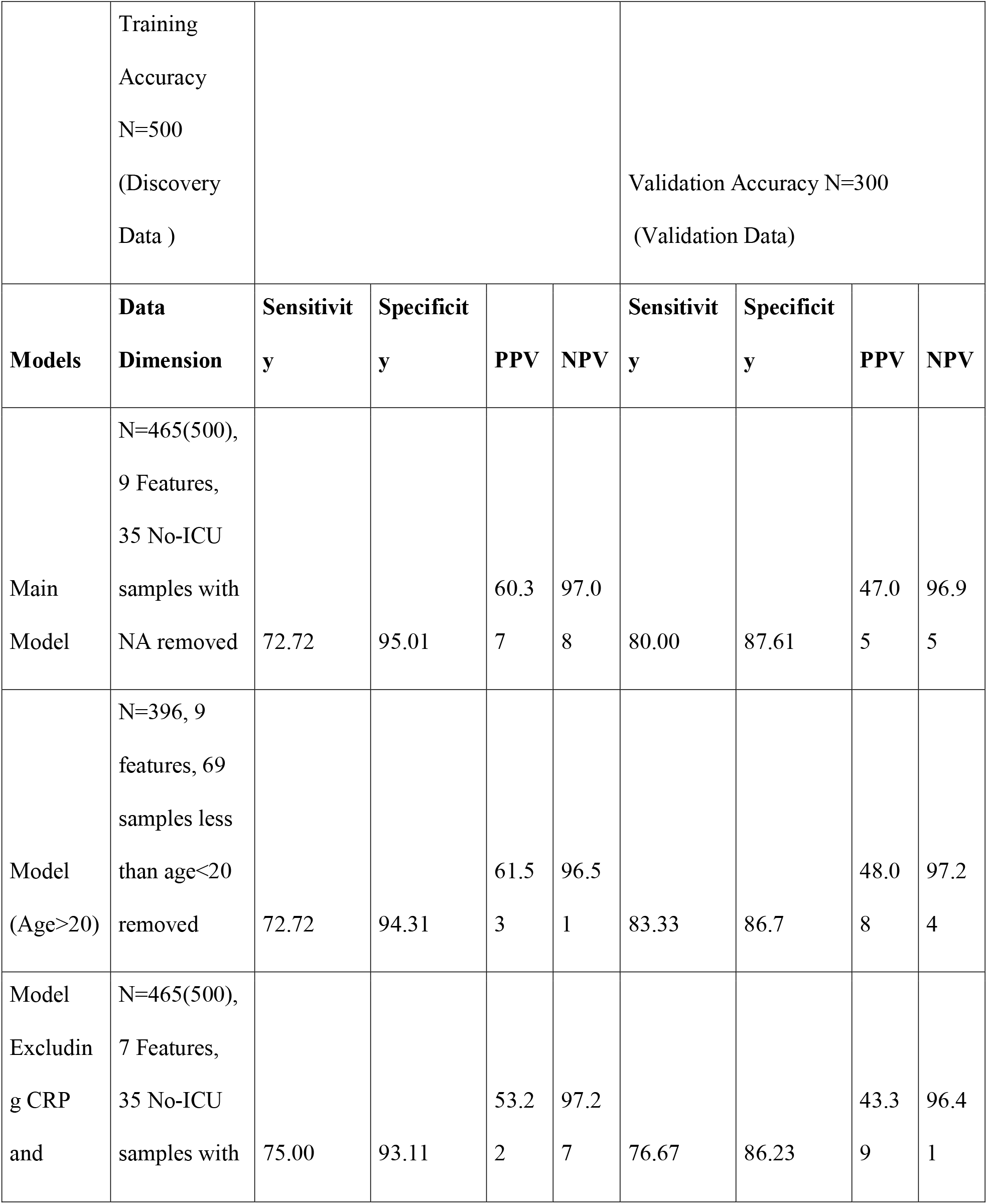
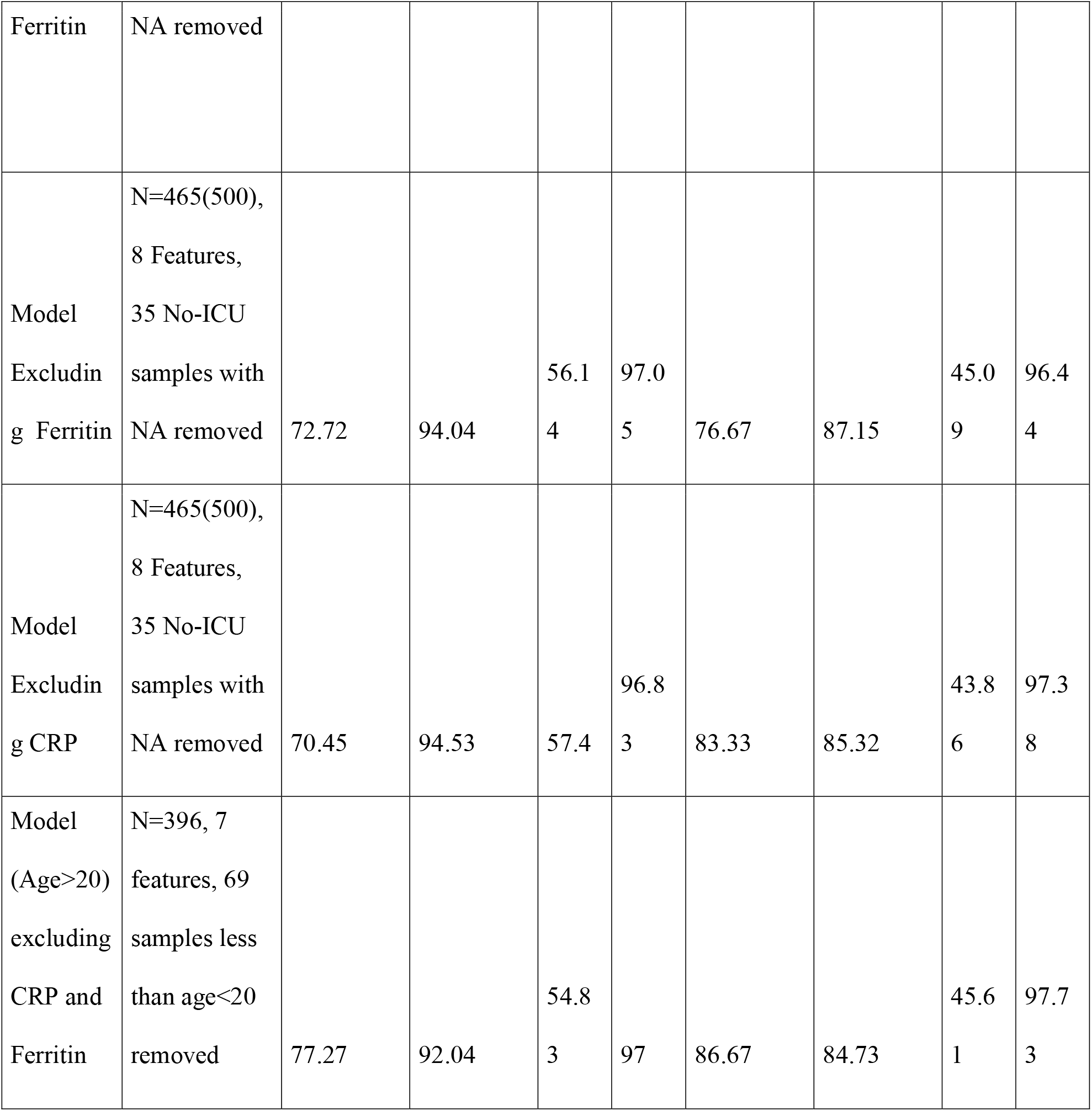
Random forest model for ICU admission, trained on first 500 patients and tested on subsequent 300patients.

Seven hundred and seventy-five patients (96.8%) were discharged from the hospital after 15-18 days of hospitalization.

## Discussion

This is one of the largest case series of COVID-19 patients from a dedicated COVID-19 hospital from Pune, India. It describes patients from the early phase of the epidemic and given a liberal hospital admission policy at the time includes a large number of asymptomatic (50%) and mild (73%) cases. Sixteen percent of patients needed ICU management, 3% died and 97% were discharged in satisfactory condition. We developed and validated a predictive model to identify patients requiring ICU admission with high accuracy using mainly clinical parameters and two commonly used markers of respiratory involvement. This supports the current policy to triage patients needing hospitalization and the usefulness of simple clinical and bed-side measurements in such a decision. This will be important in resource-limited settings.

We report on a large number of asymptomatic cases who were younger and had minimally deranged laboratory findings. Majority of them remained with mild disease and did not need any further intervention. Severe disease was associated with higher age, multiple symptoms and comorbidities, and abnormal laboratory tests.^10^ The latter included higher neutrophil (ANC) and lower lymphocyte (ALC) count leading to higher ANC/ALC ratio, and higher levels of CRP and serum ferritin concentrations, but not coagulation parameters. Mild cases had stable clinical and laboratory course while severe cases deteriorated in both. Patients who required ICU treatment were older (mean age 57 years), more symptomatic and had more comorbidities compared to those who did not require ICU treatment. Those who died were the oldest (mean age 64.7 years) and predominantly men (18/25). Our death rate of 3% is lower than the rate in Pune at that time (4.5%), probably due to preponderance of young patients with mild or asymptomatic disease.

We used both a conventional multiple logistic regression and the random forest technique to predict the need for ICU admission with high accuracy. Overriding importance of clinical and simple bedside predictors of respiratory compromise makes our model highly cost-effective in a resource limited set-up. Our results highlight the importance of having at least a basic x-ray machine in primary COVID care centers. We encourage other researchers to validate and improve our model using their data to increase its generalizability. A high negative predictive value means that the model accurately predicts those who are unlikely to need ICU admission, and raises confidence in selecting patients for home quarantine, thus helping to avoid overcrowding of specialized COVID-19 hospitals.

A large number of papers have been published on epidemiology, clinical and biochemical characters of COVID-19 patients and some models to predict serious outcomes. They are mainly from China, Italy, Spain, USA and UK.^9,10,11^ These countries have a different demographic and socio-economic profile compared to India. Their COVID-19 patients were older, more obese and with substantially more co-morbidity. This reflected in higher morbidity and mortality, especially in the geriatric and the deprived sections of the population. There are only a few reports from India: 1) Gupta et al reported on 22 young patients with mild disease from a tertiary hospital in Delhi, two-thirds having a history of travel abroad. ^12^ 2) Tambe et al reported on 197 patients (mean age 45 years) from the largest public hospital from Pune, with a high mortality rate of 29.4%, probably due to a referral bias.^13^ In other reported studies, the risk of ICU treatment varied from 8-15%.^14^ Saluja et al also reported (n=406) a profile of relatively young, predominantly male patients. Their figures of 8% ICU requirement and 1.9% mortality are not very different to our case series.^15^ We have also reported our experience of critically ill ventilated patients and outcomes. ^16^

Strengths of our study include: a large number with a mix of symptomatic and asymptomatic patients, a uniform protocol of clinical and laboratory measurements, severity classification, and ICU transfer. Inclusion of data on a large number of asymptomatic and mild patients provides an assurance that they can be managed at home or in a peripheral facility. For hospitalized patients, we developed a useful predictive model for ICU requirement from simple clinical measurements and validated it in subsequent group of patients. This has guided us to improve our treatment practices and make them pragmatic. Unfortunately, we do not have measurements of height, BMI and admission glucose concentrations in these patients, and therefore are not able to comment on their contribution to bad outcomes.

In summary, we describe clinical features of COVID-19 patients from early phase of epidemic in India and the first validated model to predict ICU requirement. This will be useful for the clinicians and the policy makers.

## Data Availability

All data is available with PI for further analysis.

## Contributors

CY, VN conceptualized this paper and drafted it with contributions from US. SS analyzed the data and other authors reviewed the study findings and contributed to the interpretation of results. All authors agreed and saw the final version of this manuscript.

## Acknowledgement

We are further grateful to all participants for giving us consent to use this data for research. **T**his research study was funded by Symbiosis International (Deemed University), Pune through its internal funds. We gratefully acknowledge the contribution of Dr Supriya Londhe, Dr Afreen Pathan, Dr Pravin Anpat, Dr Mayur Yewale, Dr Urvashi Panchal for their contribution to data collection. We thank Dr Mohan Gupte, Dr Satyajeet Rath and Dr Sanat Phatak for useful discussions and advice. The content of this paper is solely the responsibility of the authors.

