## Supplemental Tables for "Characteristics of COVID-19 patients admitted to a tertiary care hospital in Pune, India, and cost-effective predictors of intensive care treatment requirement"

**Supplementary Tables:**

Table S1: Clinical severity of COVID-19 as per MOHFW Guidelines (30^th^ March)^3^

|  | Symptoms | Clinical features |
| --- | --- | --- |
| Mild | fever, cough other generalized symptoms | pneumonia may be present  SpO2 on air >94% |
| Moderate | fever, cough, malaise +/-  breathlessness | Pneumonia +  respiratory rate 15-30/min  SpO2 90-94% on air |
| Severe | any or all | Bilateral extensive pneumonia  Respiratory Rate >30/min  SpO2 <90% on air. |

Table S2: Laboratory Investigation Protocol.

| Day1 | Day 3-5 | Day 10 |
| --- | --- | --- |
| a.Complete Blood Count(CBC) with absolute neutrophil (ANC)and lymphocyte count(ALC),  b. lactate dehydrogenase(LDH),  c. serum S. Ferritin,  d. serum C-reactive protein(CRP),  e.glucose-6-phosphate dehydrogenase(G6PD)  f. ECG and Chest x ray | a. CBC with ANC,.ALC.  b. CRP,  c. S. Ferritin,  d. Creatinine and BUN,  e. ECG and Chest X ray. | a. CBC with ANC,.ALC.  b. CRP,  c. S. Ferritin,  d. Creatinine and BUN,  e. ECG and Chest X ray. |
| D- Dimer, Sr. Procalcitonin were left to clinician discretion | If patients were put on oxygen support, shifted to HDU or ICU for further care then investigations were decided by the treating team.  D. Dimer, Sr. Procalcitonin were left to clinician discretion. | D- Dimer, Sr. Procalcitonin were left to clinician discretion |

Table S3: Demographic and clinical characteristics by sex.

| **Characteristic** | **Total**  **n (%)** | **Male**  **n (%)** | **Female**  **N (%)** | **p-value** |
| --- | --- | --- | --- | --- |
| N | 800 | 440 (55.0) | 360 (45.0) |  |
| **Age categories (y)** |  |  |  |  |
| ≤ 10y | 53 (6.6) | 35 (8.0) | 18 (5.0) | .21 |
| 10-20 | 73 (9.1) | 35 (8.0) | 18 (10.6) |  |
| 20-40 | 269 (33.6) | 150 (34.1) | 119 (33.1) |  |
| 40-60 | 250 (31.2) | 142 (32.3) | 108 (30.0) |  |
| >60y | 155 (19.4) | 78 (17.7) | 77 (21.4) |  |
| **Number of symptoms at admission** |  |  |  |  |
| Nil | 398 (49.8) | 213 (48.4) | 185 (51.4) | .35 |
| 1 | 247 (30.9) | 141 (32.0) | 106 (29.4) |  |
| 2 | 109 (13.6) | 56 (12.7) | 53 (14.7) |  |
| >2 | 46 (5.8) | 30 (6.8) | 16 (4.4) |  |
| **Symptoms** |  |  |  |  |
| Fever | 205 (25.6) | 120 (27.3) | 85 (23.6) | .24 |
| Cold-Cough | 210 (26.2) | 116 (26.1) | 94 (26.1) | .94 |
| Sore Throat | 105 (13.1) | 64 (14.5) | 41 (11.4) | .19 |
| Breathless | 90 (11.2) | 48 (10.9) | 42 (11.7) | .74 |
| Duration of symptoms (days)* | 3.4 (5.7) | 3.7 (7.5) | 3.1 (2.5) | .59 |
| **Co-morbidity** |  |  |  |  |
| T2DM | 136 (17.0) | 64 (14.5) | 72 (20.0) | .04 |
| Hypertension | 119 (14.9) | 57 (13.0) | 62 (17.2) | .09 |
| IHD/CABG | 23 (2.9) | 9 (2.0) | 14 (3.9) | .12 |
| Asthma/COPD | 13 (1.6) | 3 (0.7) | 10 (2.8) | .02 |
| **Number of comorbidity** |  |  |  |  |
| Nil | 595 (74.4) | 342 (77.7) | 253 (70.3) | .01 |
| One | 130 (16.2) | 68 (15.5) | 62 (17.2) |  |
| More than one | 75 (9.4) | 30 (6.8) | 45 (12.5) |  |
| **Severity grading at admission** |  |  |  |  |
| Mild | 587 (73.4) | 320 (72.7) | 267 (74.2) | .49 |
| Moderate | 49 (6.1) | 31 (7.0) | 18 (5.0) |  |
| Severe | 164 (20.5) | 89 (20.2) | 75 (20.8) |  |
| **Outcome** |  |  |  |  |
| Deaths | 25 (3.1) | 18 (4.1) | 7 (1.9) | .08 |
| *Values are mean (±SD) |  |  |  |  |

Table S4: ICU admitted patients’ complications and outcomes

|  | Male (n=44) | Female (n=33) | Total (n=77) |
| --- | --- | --- | --- |
| Direct ICU admission | 18 | 06 | 24 |
| Deteriorated in wards | 27 | 26 | 53 |
| Cardiac arrhythmias | 8 | 2 | 10 |
| Patients who needed invasive ventilation | 26 | 11 | 37 |
| Circulatory failure | 17 | 8 | 25 |
| AKI (KDIGO guideline) | 11 | 1 | 12 |
| Deaths | 18 | 7 | 25 |
| *Multi-system complications so will not add to 100%, values are frequency | | | |

Table S5: Demographic, clinical characteristics and comorbidity of patients by symptomatic and asymptomatic categories and by disease severity

| **Characteristic** | **Asymptomatic (n=398)^a^** | | |  | **Symptomatic (n=402) ^b^** | | |  |
| --- | --- | --- | --- | --- | --- | --- | --- | --- |
|  | **Mild** | **Moderate** | **Severe** | **P (age, sex)** | **Mild** | **Moderate** | **Severe** | **P (age, sex)** |
| N | 340 (85.4) | 17 (4.3) | 41 (10.3) | -- | 247 (61.4) | 32 (8.0) | 123 (30.6) | -- |
| Age (mean, SD) | 37.1 (19.6) | 46.0 (23.5) | 51.9 (16.5) | <.001 | 37.5 (18.4) | 56.2 (15.5) | 55.9 (15.0) | <.001 |
| **Age categories (y)** |  |  |  |  |  |  |  |  |
| ≤ 10 | 31 (9.1) | 2 (11.8) | 1 (2.4) | -- | 19 (7.7) | -- | -- | -- |
| 10-20 | 50 (14.7) | 1 (5.9) | -- |  | 21 (8.5) | -- | 1 (0.8) |  |
| 20-40 | 117 (34.4) | 5 (29.4) | 7 (17.1) |  | 114 (46.2) | 5 (15.6) | 21 (17.1) |  |
| 40-60 | 96 (28.2) | 4 (23.5) | 21 (51.2) |  | 60 (24.3) | 14 (43.8) | 55 (44.7) |  |
| >60 | 46 (13.5) | 5 (29.4) | 12 (29.3) |  | 33 (13.4) | 13 (40.6) | 46 (37.4) |  |
| **Sex** |  |  |  |  |  |  |  |  |
| Male | 185 (54.4) | 9 (52.9) | 19 (46.3) | .62 | 135 (54.7) | 22 (68.8) | 70 (56.9) | .32 |
| Female | 155 (45.6) | 8 (47.1) | 22 (53.7) |  | 112 (45.3) | 10 (31.2) | 53 (43.1) |  |
| **Symptoms** |  |  |  |  |  |  |  |  |
| Fever | -- | -- | -- | -- | 130 (52.6) | 14 (43.8) | 61 (49.6) | .59 |
| Cold-Cough | -- | -- | -- | -- | 118 (47.8) | 21 (65.6) | 71 (57.7) | .05 |
| Sore Throat | -- | -- | -- | -- | 66 (26.7) | 7 (21.9) | 32 (26.0) | .84 |
| Breathless | -- | -- | -- | -- | 16 (6.5) | 9 (28.1) | 65 (52.8) | <.001 |
| Duration of symptoms (days)* | -- | -- | -- | -- | 2.6 (1.4) | 3.2 (1.9) | 6.3 (11.4) | .004 |
| **Co-morbidity** |  |  |  |  |  |  |  |  |
| T2DM | 41 (12.1) | 6 (35.3) | 15 (36.6) | <.001 | 26 (10.5) | 5 (15.6) | 43 (35.0) | <.001 |
| Hypertension | 35 (10.3) | 3 (17.6) | 11 (26.8) | .008 | 28 (11.3) | 3 (9.4) | 39 (31.7) | <.001 |
| IHD/CABG | 5 (1.5) | -- | 1 (2.4) | -- | 5 (2.0) | 2 (6.2) | 10 (8.1) | .02 |
| Asthma/COPD | 1 (0.3) | -- | 2 (4.9) | -- | 2 (0.8) | -- | 8 (6.5) | .003 |
| **Number of comorbidity** |  |  |  |  |  |  |  |  |
| Nil | 273 (80.3) | 11 (64.7) | 22 (53.7) | <.001 | 200 (81.0) | 24 (75.0) | 65 (52.8) | <.001 |
| One | 50 (14.7) | 3 (17.6) | 11 (26.8) |  | 32 (13.0) | 7 (21.9) | 27 (22.0) |  |
| More than one | 17 (5.0_ | 3 (17.6) | 8 (19.5) |  | 15 (6.1) | 1 (3.1) | 31 (25.2) |  |
| Systolic BP | 120 (110-130) | 125 (102-134) | 120 (110-134) | .73 | 120 (110-130) | 122  (120-130) | 130 (120-140) | .86 |
| Diastolic BP | 70 (66-80) | 80 (70-90) | 75 (70-90) | .64 | 75 (70-80) | 80 (80-90) | 75 (70-80) | .02 |
| Heart rate | 78 (73-84) | 75 (71-80) | 78 (72-90) | .23 | 78 (72-84) | 79 (74-84) | 78 (69-85) | .68 |
| **Biochemical characteristics** |  |  |  |  |  |  |  |  |
| **Day 1 (n=778)** |  |  |  |  |  |  |  |  |
| Haemoglobin (g/L) | 128 (115-142) | 120 (96-137) | 123 (108-135) | .17 | 126 (112-141) | 128 (109-144) | 129 (115-140) | .73 |
| TLC (x10^3^/cmm) | 6.2 (4.8-7.6) | 6.1 (4.8-9.7) | 5.6 (4.4-7.4) | .37 | 6.0 (4.7-7.6) | 5.3  (4.8-7.1) | 7.1 (5.1-9.3) | .002 |
| Platelet count (x10^5^/cmm) | 242 (201-297) | 290 (161-368) | 231 (194-272) | .39 | 241 (184-304) | 210  (155-292) | 233 (178-304) | .16 |
| ANC (x10^3^ /cmm) | 3.2 (2.32-4.40) | 4.0 (3.05-5.75) | 3.4 (2.75-4.70) | .17 | 3.2 (2.3-4.5) | 3.5 (2.95-4.62) | 5.40 (3.20-7.50) | <.001 |
| ALC (x10^3^/cmm) | 2.0 (1.4-2.6) | 1.4 (0.9-2.3) | 1.3 (0.9-1.9) | <.001 | 1.70 (1.27-2.32) | 1.40 (1.02-1.80) | 1.10 (0.70-1.50) | <.001 |
| ANC/ALC | 1.67 (1.22-2.50) | 2.23 (1.66-6.20) | 2.34 (1.59-4.81) | <.001 | 1.91 (1.25-2.80) | 2.46 (1.80-3.54) | 4.66 (2.53-9.00) | <.001 |
| CRP (nmol/L) | 12.47 (5.42-37.61) | 50.85 (8.19-46.49) | 149.52 (30.85-591.34) | <.001 | 19.04 (5.71-67.14) | 300.00 (47.42-753.34) | 506.48 (146.09-1127.54) | <.001 |
| S. Ferritin (pmol/L) | 116.16 (35.72-260.87) | 208.07 (49.20-554.78) | 282.89 (117.51-665.56) | <.001 | 135.58 (45.88-322.12) | 533.79 (257.5-1242.09) | 810.83 (378.50-1689.60) | <.001 |
| PT (sec) | 11.1 (10.7-11.9) | 10.7 (10.3-11.2) | 12.2 (10.8-13.2) | .005 | 11.2 (10.8-11.9) | 11.8 (10.85-11.9) | 11.1 (10.6 -12.05) | .29 |
| INR | 1.01 (0.97-1.08) | 0.96 (0.94-1.01) | 1.08 (0.98-1.15) | .02 | 1.01 (0.98-1.08) | 1.07 (0.98-1.08) | 1.01 (0.96-1.09) | .28 |
| **ECG (n=356)*** |  |  |  |  |  |  |  |  |
| Normal | 145 (87.3) | 10 (90.9) | 16 (84.2) | .86 | 92 (85.2) | 10 (100.0) | 27 (64.3) | .003 |
| Abnormal | 21 (12.7) | 1 (9.1) | 3 (15.8) |  | 16 (14.8) | -- | 15 (35.7) |  |
| **Chest X-ray (800)*** |  |  |  |  |  |  |  |  |
| Normal | 326 (95.9) | 6 (35.3) | 1 (2.4) | -- | 160 (98.2) | 6 (37.5) | 3 (5.6) | <.001 |
| Abnormal | 14 (4.1) | 11 (64.7) | 40 (97.6) |  | 3 (1.8) | 10 (62.5) | 51 (94.4) |  |
| **Day 5 (n=581)** |  |  |  |  |  |  |  |  |
| Haemoglobin (g/L) | 127 (114-142) | 123 (98-134) | 123 (109-134) | .10 | 126 (110-140) | 125 (108-139) | 125 (111-138) | .77 |
| TLC (x10^3^/cmm) | 6.5 (5.4-7.6) | 6.7 (5.4-7.9) | 5.9 (5.1-6.8) | .72 | 6.30 (5.0-7.9) | 6.8 (5.32-7.35) | 8.50 (6.25-11.90) | .01 |
| Platelet count (x 10^5^/cmm) | 259.0 (208.0-309.0) | 376.5 (242.7-427.7) | 279.0 (228.0-345.5) | .03 | 259.0 (205.5-319.0) | 264.0 (166.5 -328.7) | 299.0 (220.5-396.5) | .02 |
| ANC (x10^3^/cmm) | 3.1 (2.3-4.0) | 4.05 (3.27-5.22) | 3.40 (2.50-5.00) | .16 | 3.1 (2.3-4.2) | 4.2 (3.22-4.95) | 6.30 (3.55-10.20) | <.001 |
| ALC (x10^3^/cmm) | 2.30 (1.80-2.80) | 1.50 (0.92-2.42) | 1.70 (1.10-2.20) | <.001 | 2.00 (1.50-2.60) | 1.30 (0.92-1.77) | 0.90 (0.50-1.50) | <.001 |
| ANC/ALC | 1.35 (0.96-1.92) | 2.34 (1.61-6.12) | 1.90 (1.21-4.02) | <.001 | 1.44 (1.06-2.13) | 2.85 (1.66-6.19) | 7.26 (2.49-14.41) | <.001 |
| CRP (nmol/L) | 13.14 (5.23-36.95) | 92.00(21.23-382.10) | 115.24 (14.38-396.96) | <.001 | 18.57 (6.47-68.85) | 192.38 (29.14-543.53) | 195.81 (59.23-742.96) | <.001 |
| S. Ferritin (pmol/L) | 137.96 (51.45-282.22) | 229.64 (58.19-444.23) | 333.23 (97.51-583.09) | .32 | 155.47 (65.07-375.09) | 727.19 (300.62-1222.48) | 793.19 (419.53-1859.16) | .04 |
| Creatinine (μmol/L) | 53.37 (45.75-68.62) | 61.00 (53.37-72.43) | 57.18 (45.75-70.15) | .97 | 53.37 (45.75-68.62) | 64.81 (41.93-81.58) | 68.25 (53.37-95.31) | .32 |
| **Outcome***  **ICU Transfer** | 3 (0.9) | 2 (11.8) | 6 (14.6) | -- | 2 (0.8) | 6 (18.8) | 58 (47.2) | <.001 |
| Death | 2 (0.6) | 0 | 2 (4.9) | -- | 0 | 1 (3.1) | 20 (16.3) | -- |

Note: values are Mean (SD), p-by Chi-square test; for biochemical characteristics values are Median (IQR), p by ANOVA and adjusted for age.
